# Treatment of Early Hypertension among Persons Living with HIV in Haiti: protocol for a randomized controlled trial

**DOI:** 10.1101/2021.04.13.21255408

**Authors:** Lily D Yan, Vanessa Rouzier, Eliezer Dade, Collette Guiteau, Jean Lookens Pierre, Stephano St-Preux, Miranda Metz, Suzanne Oparil, Jean William Pape, Margaret McNairy

**Affiliations:** Division of General Internal Medicine, Department of Medicine, Weill Cornell Medicine, New York, New York, USA; Center for Global Health, Department of Medicine, Weill Cornell Medicine, New York, New York, USA; Haitian Group for the Study of Kaposi’s Sarcoma and Opportunistic Infections (GHESKIO), Port-au-Prince, Haiti; Division of Cardiovascular Disease, Department of Medicine, University of Alabama at Birmingham, Birmingham, Alabama, USA

**Keywords:** Prehypertension, HIV/AIDS, low-middle income country, Haiti, clinical trial, amlodipine

## Abstract

**Background:** People living with HIV (PLWH) are at increased risk of cardiovascular disease (CVD) and death, with greater burdens of both HIV and CVD in lower-middle income countries. Treating prehypertension in PLWH may reduce progression to hypertension, CVD risk and potentially mortality. However, no trial has evaluated earlier blood pressure treatment for PLWH. We propose a randomized controlled trial to assess the feasibility, benefits, and risks of initiating antihypertensive treatment among PLWH with prehypertension, comparing prehypertension treatment to standard of care following current WHO guidelines.

**Methods:** A total of 250 adults 18-65 years and living with HIV (PLWH) with viral suppression in the past 12 months, who have prehypertension will be randomized to prehypertension treatment versus standard of care. Prehypertension is defined as having a systolic blood pressure (SBP) 120-139 mmHg or diastolic blood pressure (DBP) 80-89 mmHg. In the prehypertension treatment arm, participants will initiate amlodipine 5 mg daily immediately. In the standard of care arm, participants will initiate amlodipine only if they develop hypertension defined as SBP ≥ 140 mmHg or DBP ≥ 90 mmHg. The primary outcome is the difference in mean change of SBP from enrollment to 12 months. Secondary outcomes include feasibility, acceptability, adverse effects, HIV viral suppression, and medication adherence. Qualitative in-depth interviews with providers and participants will explore attitudes about initiating amlodipine, satisfaction, perceived CVD risk, and implementation challenges.

**Discussion:** PLWH have a higher CVD risk and may benefit from a lower BP threshold for initiation of antihypertensive treatment.

**Trial registration:** Clinicaltrials.gov registration number NCT04692467, registration date December 15, 2020, protocol ID 20-03021735.

## Background

People living with HIV (PLWH) are at an increased risk of cardiovascular disease (CVD), including hypertension, myocardial infarction (MI) and stroke due to a complex interplay between increased inflammation from HIV, adverse effects of antiretroviral therapy (ART), and traditional host risk factors (1). Hypertension prevalence is 35% for PLWH on ART, and the relative risk for MI and stroke is as much as two-fold higher in PLWH compared to people without HIV (2–6).

The highest burden of HIV has long been in low- and middle-income countries (LMICs) (7). The majority of persons with hypertension are now also located in LMICs—an estimated 1.04 billion—compared to 349 million in high-income countries (HIC) (8). High systolic blood pressure (SBP) has become the leading risk factor for all-cause mortality in the world over the past 30 years (9). Haiti exemplifies this dual burden, with the highest HIV prevalence in the western hemisphere (10), and an age-standardized hypertension prevalence of 29% (11,12). Our prior research has shown that among PLWH in Haiti, hypertension is independently associated with increased mortality (HR 2.47 [95% CI 1.10-5.57]), after adjusting for immune status, age, and sex (13).

Given the step-wise increased risk of CVD events and mortality among people with elevated SBP > 115 mmHg (14,15), there may be significant benefits to treating prehypertension, defined as SBP 120-139 mmHg or diastolic blood pressure (DBP) 80-89 mmHg. Treatment of prehypertension not only prevents progression to hypertension (16), it can also decrease CVD events and disease progression in people with diabetes, chronic kidney disease (CKD), and nonobstructive coronary artery disease (17–20).

Haiti guidelines for hypertension treatment follow the World Health Organization (WHO) guidelines, which recommend initiating antihypertensive treatment at SBP ≥140 mmHg or DBP ≥90 mmHg for the general adult population, and at SBP ≥130 mmHg or DBP ≥80 mmHg for high risk groups such as people with diabetes or CKD (21). In contrast, the 2017 American College of Cardiology / American Heart Association (ACC/AHA) guidelines recommend treatment initiation at SBP ≥140 mmHg or DBP ≥90 mmHg for people with 10 year predicted CVD risk < 10%, and at SBP ≥130 mmHg or DBP ≥80 mmHg for people with 10 year predicted CVD risk of ≥ 10% (22). The WHO does not include PLWH in their definition of high-risk groups despite PLWH having a similarly increased CVD risk compared to people with diabetes or CKD (1). The AHA/ACC guidelines also do not adequately capture risk in PLWH given traditional 10 year CVD risk prediction models underestimate risk for PLWH (23).

The Treatment of Early Hypertension among Persons Living with HIV randomized controlled trial aims to assess the feasibility, benefits, and risks of initiating antihypertensive treatment among PLWH with prehypertension, comparing prehypertension treatment to the WHO standard of care in parallel groups. These data will inform a future large trial, designed with sufficient power to detect clinically significant differences in BP, and differences in rates of CVD events and mortality, between a prehypertension treatment versus standard of care arm among PLWH.

## Methods

### Study design and site

This is an unblinded randomized controlled trial of 250 PLWH, with half randomized to the prehypertension treatment arm, and half to the standard of care. All participants have prehypertension, defined as SBP 120-139 mmHg or DBP 80-89 mmHg at two separate clinic visits. In the prehypertension treatment arm, participants will be started on amlodipine 5mg daily at enrollment. In the standard of care arm, participants will not be started on amlodipine at enrollment.

The study site is the Groupe Haitien d’Etude du Sarcome de Kaposi et des Infections Opportunistes clinics (GHESKIO) in Port-au-Prince, Haiti. GHESKIO is a medical facility that has operated continuously over the past 40 years conducting clinical care, research, and advocacy for PLWH and associated chronic diseases with over 600,000 patient visits annually. This study will recruit patients from the GHESKIO’s Adult HIV Clinic, which cares for over 175,000 patients annually with HIV.

### Study population

The study population will include 250 PLWH receiving HIV care at GHESKIO. People aged 18-65 years of age on ART for one year, with HIV viral load suppression defined as < 1000 copies/ml in the past 12 months, and with confirmed prehypertension (SBP 120-139 mmHg or DBP 80-89 mmHg) not currently on antihypertensives will be included (Table 1). Younger PLWH are intentionally included as hypertension prevalence among young adults in Haiti is up to three-fold higher (10,11) than among age-matched African Americans in US cohorts (24). Participants with CKD or diabetes are excluded as Haitian guidelines initiate treatment at a threshold of SBP ≥ 130 mmHg or DBP ≥ 80 mmHg. PLWH on a protease inhibitor such as ritonavir are excluded because of possible drug-drug interactions with amlodipine (25).

**Table 1:** Inclusion and exclusion criteria for study population.

| Inclusion criteria | Exclusion criteria |
| --- | --- |
| PLWH 18-65 years of age | Pregnancy |
| ART duration ≥ 1 year, stable regimen ≥ 6 months | Chronic kidney disease or diabetes |
| Viral suppression: HIV 1-RNA < 1,000 copies/mL within past 12 months | On protease inhibitor |
| Prehypertension:<br>SBP 120-139 and DBP < 90, OR<br>SBP < 140 and DBP 80-89 mmHg | Advanced illness with limited life expectancy |
| Not currently on antihypertensive medications | Plans to move out of the area within the next year |
| Receives HIV care at GHESKIO | Clinician determination that patient is unstable on ART |
| Willing to provide consent |  |

### Intervention

The study intervention in the prehypertension treatment arm is initiation of amlodipine 5mg daily immediately. The study intervention in the standard of care arm is initiation of amlodipine 5mg daily only if participants develop incident hypertension (SBP ≥ 140 mmHg or DBP ≥ 90 mmHg) during the course of the study, as recommended by Haiti’s hypertension treatment guidelines (21,26). Throughout the follow-up period, if a participant randomized to the prehypertension treatment arm has a SBP ≥ 130 mmHg after 1 month of treatment, amlodipine will be increased from 5mg to 10mg. If a participant develops new or changing symptoms, he/she will be referred to GHESKIO’s CVD clinic for medical care by clinic staff.

Amlodipine discontinuation or dose decrease will be based on development of adverse effects or participant request, after review by an independent Drug and Safety Monitoring Board composed of external experts in cardiovascular clinical research and biostatistics. Adverse events include dizziness, fainting, and lower extremity edema. Only Grade III-V adverse effects will trigger amlodipine discontinuation, modeled after the NIH Division of AIDS (DAIDS) Table for Grading Severity of Adult and Pediatric Adverse Events (27).

### Study visits and measurements

There are four categories of study visits: recruitment and screening, enrollment, follow-up, and a final 12-month visit (Figure 1).

**Figure 1:**
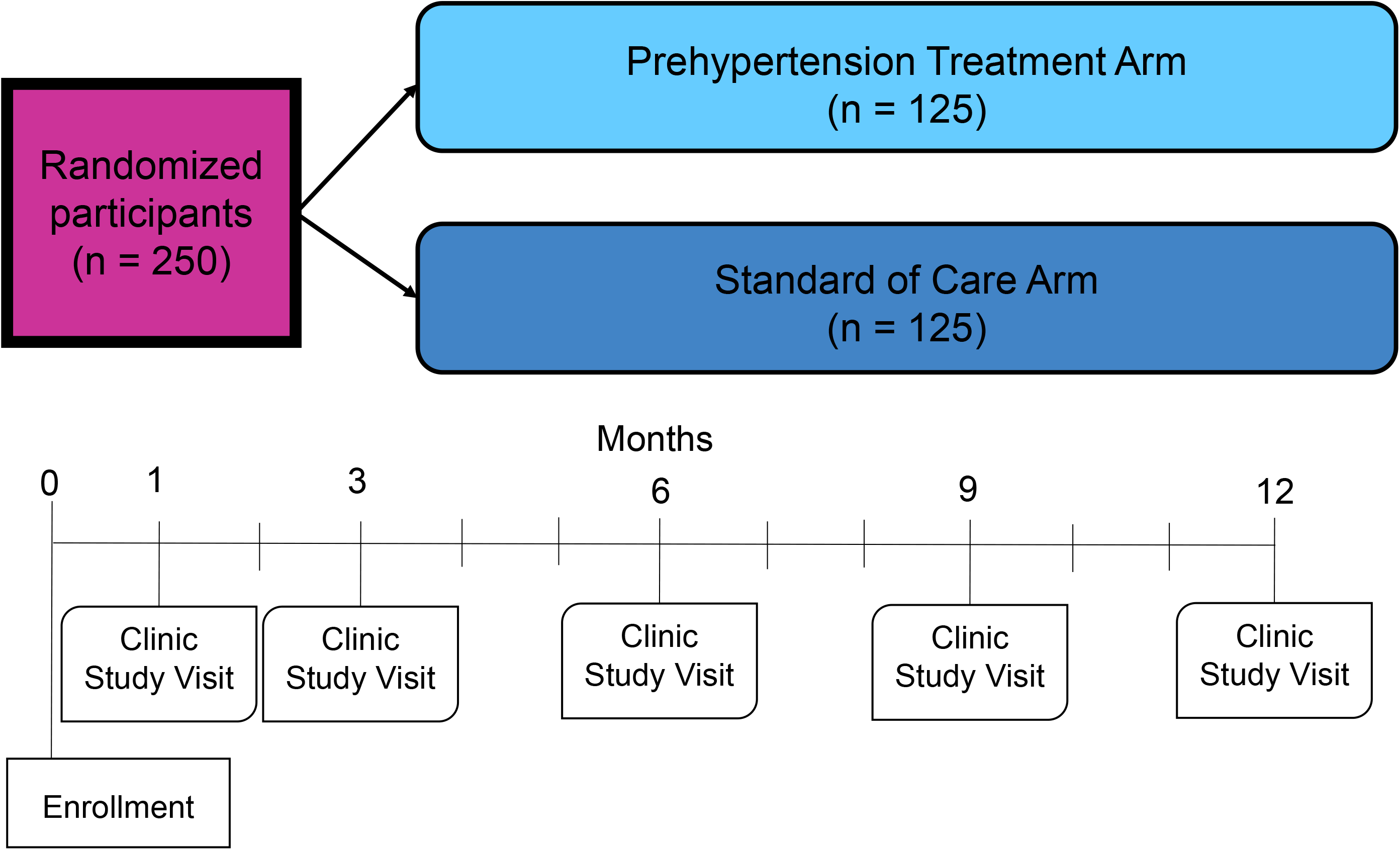
Study Overview. Legend: Study follow-up visits will occur in the GHESKIO Clinic, with community visits to measure BP offered in months 2, 5, and 8.

#### Recruitment, screening and informed consent

Participant recruitment will occur at the GHESKIO Adult HIV clinic as well as through a review of the GHESKIO electronic medical record to pre-identify PLWH who meet inclusion criteria. Individuals will then be invited for a screening visit and informed consent.

In the first part of the screening visit, study eligibility criteria will be evaluated and blood pressure (BP) will be measured. Two BP measurements, on different days, in the prehypertensive range are needed to qualify for the study.

All BP measurements will follow ACC/AHA and WHO international guidelines (21,22). We will use semi-automated electronic cuffs (OMRON 742-5 series in the community, OMRON HEM 907 in clinic). After resting for 5 minutes, the participant will have three BPs measured, separated by 1-minute intervals. The average of the 3 BPs is the BP for the study visit.

Individuals found to have at least two BPs in the prehypertensive range will be introduced to the study by research staff, and if interested in participating, will provide written informed consent. Subsequent screening procedures for consented participants include screening labs for pregnancy, diabetes, and CKD (Table 2). Those without evidence of pregnancy, diabetes and or renal disease will proceed with randomization.

**Table 2:** Study Measures and Schedule.

| TIMEPOINT | SCREENING VISIT | ENROLLMENT | M1 | M2* | M3 | M5* | M6 | M8* | M9 | M12 |
| --- | --- | --- | --- | --- | --- | --- | --- | --- | --- | --- |
| **SCREENING** |  |  |  |  |  |  |  |  |  |  |
| **Screening Labs:** urine/serum pregnancy test, fasting or random glucose, creatinine | X |  |  |  |  |  |  |  |  |  |
| **ENROLLMENT** |  |  |  |  |  |  |  |  |  |  |
| **Demographics:** age, sex |  | X |  |  |  |  |  |  |  |  |
| **Socioeconomic factors:** income, education, occupation, marital status, children. |  | X |  |  |  |  |  |  |  |  |
| **Medical History:** history of CVD risk factors and diseases HIV, tuberculosis |  | X |  |  |  |  |  |  |  |  |
| **Family History:** stroke, myocardial infarction, or heart failure in parents and 4 oldest siblings |  | X |  |  |  |  |  |  |  |  |
| **Health behaviors:** current cigarette smoking, alcohol, physical activity |  | X |  |  |  |  |  |  |  | X |
| **Multidimensional Poverty:** child death, educational attainment, household construction materials, possessions |  | X |  |  |  |  |  |  |  |  |
| **Allocation** |  | X |  |  |  |  |  |  |  |  |
| **INTERVENTIONS** |  |  |  |  |  |  |  |  |  |  |
| **Treatment of Prehypertension** |  | X | X | X | X | X | X | X | X | X |
| **Standard of Care** |  | X | X | X | X | X | X | X | X | X |
| **ASSESSMENTS** |  |  |  |  |  |  |  |  |  |  |
| **Physiologic:** height, weight |  | X |  |  |  |  |  |  |  | X |
| **Blood pressure** |  | X | X | X | X | X | X | X | X | X |
| **ECG** |  | X |  |  |  |  |  |  |  |  |
| **Echocardiography and vascular ultrasound** |  | X |  |  |  |  |  |  |  |  |
| **Enrollment Labs:** total cholesterol, HDL, CD4, HIV-1 RNA viral load |  | X |  |  |  |  |  |  |  |  |
| **12M Labs:** HIV-1 RNA viral load |  |  |  |  |  |  |  |  |  | X |
| **Medical record abstraction:** diagnoses codes (ICD-9), laboratory measures, diagnostic imaging, and cause of death among participants who receive clinical care from the GHESKIO NCD clinic, a GHESKIO-affiliated hospital, or other health facility |  |  | X | X | X | X | X | X | X | X |
| **Adverse Events:** if in treatment group |  |  | X | X | X | X | X | X | X | X |
| **In depth Interviews** |  |  |  |  |  |  |  |  |  | X |
| **Medication Adherence (ART and amlodipine)** |  | X | X | X | X | X | X | X | X | X |

Legend: \* indicates a study visit in the community.

#### Enrollment procedures

The study enrollment visit includes a questionnaire, physical exam, and laboratory tests. The questionnaire includes demographic and socioeconomic information and information about cardiovascular disease health behaviors using validated instruments on smoking (36), diet (36), alcohol use (37), and physical activity (38). Medical history of cardiovascular risk factors, HIV, tuberculosis, and family history of cardiovascular disease will be ascertained. Vital signs, including BP, will be obtained and a physical exam will be performed. Laboratory data including total cholesterol, high density lipoprotein (HDL), CD4 cell count, and HIV-1 RNA viral load will be collected. Lastly, an EKG and echocardiogram will be performed.

Patients will be randomized to the 1) prehypertension treatment arm, or 2) standard of care arm in blocks of 10 using a random number generator by the GHESKIO Clinical Trial Unit’s computer algorithm.

#### Follow-up study visits

Participants will be evaluated at five study visits during the 12-month follow-up at GHESKIO (Figure 1). The first will occur within 1 month of enrollment to evaluate response to initiation of amlodipine (if applicable). Subsequent visits will occur in months 3, 6, 9 and 12. Each will include BP measurement, evaluation for medication adherence, drug side effects, and inquiry about interim hospitalizations. Research staff will review outside hospitalizations for new CVD-related diagnoses and events. Every visit will include lifestyle counseling on diet, exercise, and general medication adherence. ART adherence will be measured using validated AACTG Adherence Instruments (28), and amlodipine adherence will be measured using the Hill-Bone instrument (29). At the 12-month visit, a physical exam, study questionnaire about health behaviors, and a HIV-1 RNA viral load assessment will be performed. ART will be distributed in 6-month supply per national guidelines in both study arms. Amlodipine will be distributed in 3-month supply after the first follow-up visit.

In addition, follow-up visits at months 2, 5, and 8 will take place in the community at the location of participant preference. These visits will include the same assessments.

#### In-depth interviews

In-depth interviews will be conducted by study healthcare providers (physicians, nurses) in a subset of 30 participants in the prehypertension treatment arm. We will explore attitudes about initiating amlodipine in addition to existing HIV medications, positive and negative perceptions about amlodipine, satisfaction with treatment, intent to continue treatment, home BP measurement, and perceived CVD risk. We will interview providers about implementation challenges and unintended consequences. Interviews will be conducted in Creole, audio-recorded, transcribed verbatim, and translated into English for analysis using Atlas-ti.

All study data will be collected on a secure web platform using REDCap (supported by the National Center for Advancing Translation Science of the National Institute of Health under award number UL1TR002384).

### Study outcomes and measures

The primary outcome is the change in mean SBP (mmHg) from enrollment to 12 months and the difference in this change between the two study arms. Secondary outcomes include feasibility, acceptability, adverse effects, HIV viral suppression at 12 months, and ART medication adherence. Feasibility will be measured as the percent of participants retained between enrollment and 12 months, across both study arms and adherence to amlodipine using a validated scale (29). Acceptability will be measured through key themes that emerge from semi-structured key informant qualitative interviews with participants and providers (e.g. perceived benefits of amlodipine, perceived harms). Adverse effects will be measured as number of subjects who have adverse events related to amlodipine, including dizziness, fainting, lower extremity edema, and any other symptoms which may be related. Changes in HIV viral suppression will be measured as the change in number of participants with HIV-1 RNA viral loads < 1,000 copies/mL from enrollment to 12 months. HIV medication adherence will be measured by number of participants with > 90% adherence using 4 day pill recalls (28).

### Power and sample size calculations

With a sample of 250 participants across both arms and 8 time points for BP measurements for each participant, we have 80% power with alpha 0.05 to detect a difference in change in SBP of 4 mmHg or more between the study arms at 12 months (assuming consistent difference across time and a conservative correlation among measurement of 0.95, standard deviation of BP at each time point = 25 mmHg) (30). We will present summary statistics (e.g., mean and SD of pre, post, and difference within and between arms).

### Statistical methods

For the primary outcome, we will compare the mean difference in change in SBP among participants from enrollment to 12 months in each study arm using linear mixed-effects model (LMM) accounting for repeated measures and correlations within subjects, where the time*treatment interaction will serve as the “primary” parameter addressing differential slopes for the two arms. We anticipate that the two arms will have similar baseline characteristics due to randomization, and therefore the primary analysis will not adjust for baseline variables. If baseline variables are imbalanced, we will adjust factors in regression and report the findings as a sensitivity analysis. We will assess the incidence of HTN using Kaplan Meier methods. We will analyze dichotomized outcomes (SBP <120 and DBP <80) at all time points via a generalized mixed-effects model and at 12 months via Fisher exact tests and logistic regression.

In other sensitivity analyses, we will explore the following issues: 1) compliance/adherence (per protocol analysis); 2) missing data/dropout (via last-value-carried-forward, multiple imputation or inverse-probability weighting); 3) competing risks (e.g., death); 4) joint modeling (longitudinal and survival data); and 5) changepoint (treatment initiation in standard of care arm).

For secondary outcomes, we will use a range of analytic methods. Feasibility will be measured as the proportion of eligible participants willing to participate in the trial and their retention and the proportion initiating amlodipine. The proportion of participants in each study arm with viral suppression (HIV-1 RNA < 1000 copies/mL) at 12 months will be compared between study arms using the Fisher exact test. We assume that participants who are lost to follow-up or dead at 12 months will have HIV-1 RNA > 1000 copies/mL. Other categorical data including medication adherence, CVD risk factors (e.g. diabetes) will be analyzed similarly.

Longitudinal data such as adherence and adverse events (continuous or binary/categorical) will be analyzed accounting for repeated measures within participants, via LMM and generalized LMM as described above. We will also consider the generalized estimating equation-based method (so called, marginal or population-averaged model) and will report consistency or meaningful discrepancy. Acceptability will be assessed in qualitative interviews by trained research staff using interview information transcribed verbatim, translated into English, and then coded for analysis using grounded theory.

### Ethics and dissemination

This study was approved by institutional review boards (IRBs) at Weill Cornell Medicine and GHESKIO. Written informed consent by participants will be obtained by trained study personnel using existing protocols in the GHESKIO Clinical Trial Unit, including testing for comprehension on study intervention and measurements.

Important protocol modifications will be shared with investigators and trial participants within two weeks, and submitted to IRBs and the trial registry within 30 days. Trial results will be communicated to participants and local healthcare providers through a series of web-based and in-person presentations. Results will be communicated to the public via publications and the clinicaltrials.gov registry.

## Discussion

The purpose of the Treatment of Early Hypertension among Persons Living with HIV randomized controlled trial is to assess the feasibility, benefits, and risks of initiating antihypertensive treatment among PLWH with prehypertension. Treating prehypertension may reduce progression to hypertension and reduce CVD risk and potentially CVD death in PLWH, and no trial has evaluated use of a lower BP treatment threshold of SBP ≥ 130 or DBP ≥ 80 for this specific population. This trial is a first step towards a large randomized controlled trial adequately powered for clinical outcomes, including CVD, to evaluate the effects of prehypertension treatment in PLWH.

Prior research clearly links both hypertension and prehypertension to higher CVD risk and death in PLWH. In the Veterans Aging Cohort Study Virtual Cohort, having HIV and prehypertensive BP was associated with an increased risk of acute myocardial infarction compared to veterans without HIV, with low prehypertensive BPs (SBP 120-129, DBP 80-84) HR 1.60 [95% CI 1.07-2.39] and high prehypertensive BPs (SBP 130-139, DBP 85-89) HR 1.81 [95% CI, 1.22-2.68] (3). In Haiti, our prior research has shown that hypertension in PLWH at the time of ART initiation is associated with increased mortality. In a cohort of 816 PLWH who started ART in 2005-2008 at GHESKIO, 5.3% had hypertension at the time of ART initiation, and this independently predicted mortality during 10 years of follow-up (HR 2.47 [95% CI 1.10-5.57]), after adjusting for age, sex, CD4 count at ART start (13). One third of the deaths were from stroke.

This trial is designed to help fill in knowledge gaps including: Is it feasible to initiate and integrate antihypertensive treatment into HIV care among a relatively healthy cohort of PLWH with prehypertension? Could initiating first-line antihypertensive medication worsen HIV outcomes? What is the BP reduction using amlodipine in PLWH? Data from this trial will inform the design of a larger randomized trial powered for incident CVD events.

Our long-term goal is to change guidelines to institute earlier initiation of antihypertensive medication in PLWH with prehypertension in order to prevent CVD in this population. The 2017 ACC/AHA guidelines recommend using a combination of absolute CVD risk and BP level to guide treatment. However, traditional 10 year CVD risk prediction models underestimate risk for PLWH, with poor to moderate discrimination (c statistics 0.65 to 0.73), and higher observed versus predicted risk at all risk levels (31). These guidelines may also be unduly burdensome to implement in low-middle income countries, where laboratory tests for cholesterol may not be widely available or affordable. The SPRINT trial evaluated initiating antihypertensive treatment to a goal of SBP ≤ 120 mmHg among people with increased cardiovascular risk, and found that intensive treatment resulted in fewer fatal and non-fatal CVD events and all-cause mortality (20). Accurately capturing the increased CVD risk among PLWH and translating it into an actionable treatment threshold is important, and one way to achieve this may be to enforce a lower SBP treatment target.

Strengths of this study include a randomized controlled trial design to equally distribute covariates and avoid omitted variable bias. Amlodipine is a relatively safe blood pressure medication, and requires no lab testing for monitoring (unlike hydrochlorothiazide which requires checking blood electrolytes and renal function). Frequent follow-up visits will closely monitor for hypotension and other adverse effects of amlodipine, as well as track a possible impact on ART adherence. We include community-based BP measurement which will allow assessment of BP in non-clinic settings. The study builds upon the strong infrastructure of the HIV Clinic at GHESKIO, expanding its focus on CVD among PLWH. This will be among the first studies evaluating BP in PLWH treated with dolutegravir, an ART associated with weight gain that became the first-line regimen in Haiti and many other LMICs since 2018. Finally, we include echocardiographic assessment of pre-existing myocardial and vascular dysfunction among PLWH, an entity that has not been well described in the past. Our sample size of 250 participants is not powered for CVD events.

## Conclusion

There is an urgent need for CVD prevention among PLWH with elevated BP who have an alarmingly high risk of CVD events and mortality. This study is the first step toward evaluating the feasibility, benefit, and safety of earlier hypertension treatment of PLWH. Our data will inform a larger trial powered for CVD events that could change the paradigm for hypertension treatment among PLWH.

## Supporting information

SPIRIT checklist

## Data Availability

The datasets to be collected during the current study will be available from the corresponding author on reasonable request. Data request should be submitted to Dr. Margaret McNairy who will review the data request with Haiti GHESKIO Site PI, Dr. Jean Pape and the study's Data Safety Monitoring Board for approval.

## Declarations

### Ethics approval and consent to participate

This study was approved by institutional review boards at Weill Cornell Medicine and GHESKIO.

### Consent for publication

Not applicable

### Authors contributions

Conceived study: VR, JWP, MLM

Data curation: N/A

Formal analysis: N/A

Funding acquisition: MLM

Investigation: VR, ED, CG, JWP

Methodology: LDY, VR, ED, CG, SS, MM, SO, JWP, MLM

Project administration and resources: VR, ED, CG, SS, JWP

Software: LDY

Writing-original draft preparation: LDY, MLM

Writing-review & editing: LDY, VR, ED, SS, MM, SO, JWP, MLM

All authors have read, and confirm that they meet, ICMJE criteria for authorship.

## Acknowledgements

We acknowledge the valuable input from the Haitian College of Cardiology, the community health workers who help with data collection, and the patients at GHESKIO for entrusting us with their care.

